# Determinants of SARS-CoV-2 nasopharyngeal testing in a rural community sample susceptible of first infection: the CHRIS COVID-19 study

**DOI:** 10.1101/2022.01.27.22269941

**Authors:** Daniele Giardiello, Roberto Melotti, Giulia Barbieri, Martin Gögele, Christian X. Weichenberger, Luisa Foco, Daniele Bottigliengo, Laura Barin, Rebecca Lundin, Peter P. Pramstaller, Cristian Pattaro

## Abstract

To characterize COVID-19 epidemiology, numerous population-based studies have been undertaken to model the risk of SARS-CoV-2 infection. Less is known about what may drive the probability to undergo testing. Understanding how much testing is driven by contextual or individual conditions is important to delineate the role of individual behavior and to shape public health interventions and resource allocation. In the Val Venosta/Vinschgau district (South Tyrol, Italy), we conducted a population-representative longitudinal study on 697 individuals susceptible to first infection who completed 4,512 repeated online questionnaires at four week intervals between September 2020 and May 2021. Mixed-effects logistic regression models were fitted to investigate associations of self-reported SARS-CoV-2 testing with individual characteristics (social, demographic, and biological) and contextual determinants. Testing was associated with month of reporting, reflecting the timing of both the pandemic intensity and public health interventions, COVID-19-related symptoms (odds ratio, OR:8.26; 95% confidence interval, CI:6.04–11.31), contacts with infected individuals within home (OR:7.47, 95%CI:3.81–14.62) or outside home (OR:9.87, 95%CI:5.78–16.85), and being retired (OR:0.50, 95%CI:0.34-0.73). Symptoms and next within- and outside-home contacts were the leading determinants of swab testing predisposition in the most acute phase of the pandemics. Testing was not associated with age, sex, education, comorbidities, or lifestyle factors. In the study area, contextual determinants reflecting the course of the pandemic were predominant compared to individual sociodemographic characteristics in explaining the SARS-CoV-2 probability of testing. Decision makers should evaluate whether the intended target groups were correctly prioritized by the testing campaign.

## Introduction

The coronavirus disease 2019 (COVID-19) is severely threatening global health, with more than 650 million affected individuals and nearly 7 million deaths worldwide by December 2022 (1). Population-based studies are central to assess incidence and identify determinants of infection (2–4), disease severity, and mortality (5–9).

Although many such studies have been conducted, few have had a joint focus on contextual and individual determinants of SARS-CoV-2 testing, despite evidence that personal characteristics can impact health behaviors. For instance, healthcare workers had a higher risk of SARS-CoV-2 infection compared to the general population (10), but they were also periodically tested to minimize the spread of infection among hospitalized individuals (11). Differential access to SARS-CoV-2 testing by educational level and income has also been observed (12,13). In other contexts, financial barriers were not associated with undergoing a test (14). Many other individual determinants of testing are currently unexplored. For instance, females exhibit generally more health-prone behaviors than males (15) but the probability to undergo a SARS-CoV-2 test seems to be context-dependent with, for instance, female healthcare workers who may be tested less than their male colleagues (16). Observational studies suggest an association between SARS-CoV-2 infection incidence and sex (2,3,17,18). Whether this association is mainly biological in nature (19) or also driven by differential behaviours by sex is an open question. Similar reasoning applies to other determinants already associated with SARS-CoV-2 infection, whether biological such as age (20) or sociocultural such as educational level, employment (11), lifestyle, and income (2). Concerns about the pandemic and knowing someone with a diagnosis of COVID-19 were also associated with more testing (14). Other contextual determinants of testing may include temporal variability of testing capacity and resources availability, which depend not only on healthcare system organisation but also on the pandemic evolution.

We studied the case of the Val Venosta/Vinschgau district in South Tyrol (Italy) between September 2020 and May 2021. Until the end of summer 2020, the district was nearly free of SARS-CoV-2 infections. With the start of the school season and the advent of autumn, there was a rapid increase of infections. By the end of October 2020, a number of municipalities were defined as red zones in which testing was being stepped up to track and contain infections (21). Following an explosive increase in infections, South Tyrol conducted mass testing using rapid antigen tests, involving 70% of the whole population between November 18^th^ and 25^th^, 2020 (22). Meanwhile, the availability of rapid nasal tests in pharmacies and healthcare facilities became widespread, with a substantial increase in the effective testing capacity and easier access to testing for the population. At the beginning of 2021, the vaccination campaign began, starting with the most exposed and vulnerable population groups. In February and March 2021 there was a new wave of infections and hospitalisations, with a reduction of infections by late spring.

Understanding to which extent COVID-19 testing probability may be driven by general or personal conditions is key to evaluate public health interventions and resource allocation. To assess this question we exploited data collected in the context of a longitudinal population-based study on COVID-19 conducted in this rural Alpine region and characterized by repeated surveys to study participants every four weeks for the entire duration of the study (23). All participants included in this analysis were negative to previous SARS-CoV-2 infection and thus susceptible to first infection. Individual determinants comprised biological, health, behavioral, cultural, and socio-economic aspects. The pandemic pattern was captured by the month at follow-up participation. Results for the probability of SARS-CoV-2 testing are presented according to the relative contribution of both the epidemic pattern and individual characteristics.

## Material and Methods

### Study design

The CHRIS COVID-19 study (23), a longitudinal study embedded within the Cooperative Health Research In South Tyrol (CHRIS) study (24), was established on July 13, 2020 to assess the determinants of SARS-CoV-2 infection and COVID-19 disease. The study was approved by the Ethics Committee of the Healthcare System of the Autonomous Province of Bolzano/Bozen (deliberation number 53-2020). All participants provided informed consent.

As extensively described elsewhere (23), we estimated that a stratified random sample of 1450 individuals, derived from all the 13,393 CHRIS participants to represent the general adult population of the Val Venosta/Vinschgau district based on age and sex strata distribution, would suffice to estimate a cumulative incidence between 0.01% and 1.1% with a 99% confidence level. To achieve this goal we oversampled 1812 individuals, assuming a 80% participation rate.

By August 28, 2020, all selected individuals were invited to a baseline assessment consisting of a Roche Elecsys® Anti-SARS-CoV-2 assay serum antibody (SAb) test, a swab real time polymerase chain reaction (PCR) test, and a screening questionnaire. This baseline questionnaire referred to the period from February 1, 2020 until the participation date and included questions on sociodemographic and dwelling information, lifestyle, regular medication, past and current health status, and a section dedicated to SARS-CoV-2 infection: previous diagnosis, symptoms, within- or outside-home contacts with infected or symptomatic individuals, and vaccination.

Follow-up of those with negative baseline tests was conducted by administering a streamlined online questionnaire every 4 weeks for one year, limited to SARS-CoV-2 relevant information and incident events, including symptoms, testing, infection, vaccination, and recent contacts at risk. Participants reporting a positive a naso/oropharyngeal swab test as PCR, antigen test or saliva swab test (hereafter: ‘testing’) or serological test or a contact with a positive individual since the last interview were invited to the study center for PCR and SAb testing. If positive to any of the tests at the study center, the participant was excluded by design from further follow-up survey questionnaires as they were involved in a separate longitudinal evaluation of antibody response. If negative to both tests, the participant was considered still susceptible to infection and eligible for the follow-up questionnaires every four weeks (**Figure 1**).

**Figure 1.**
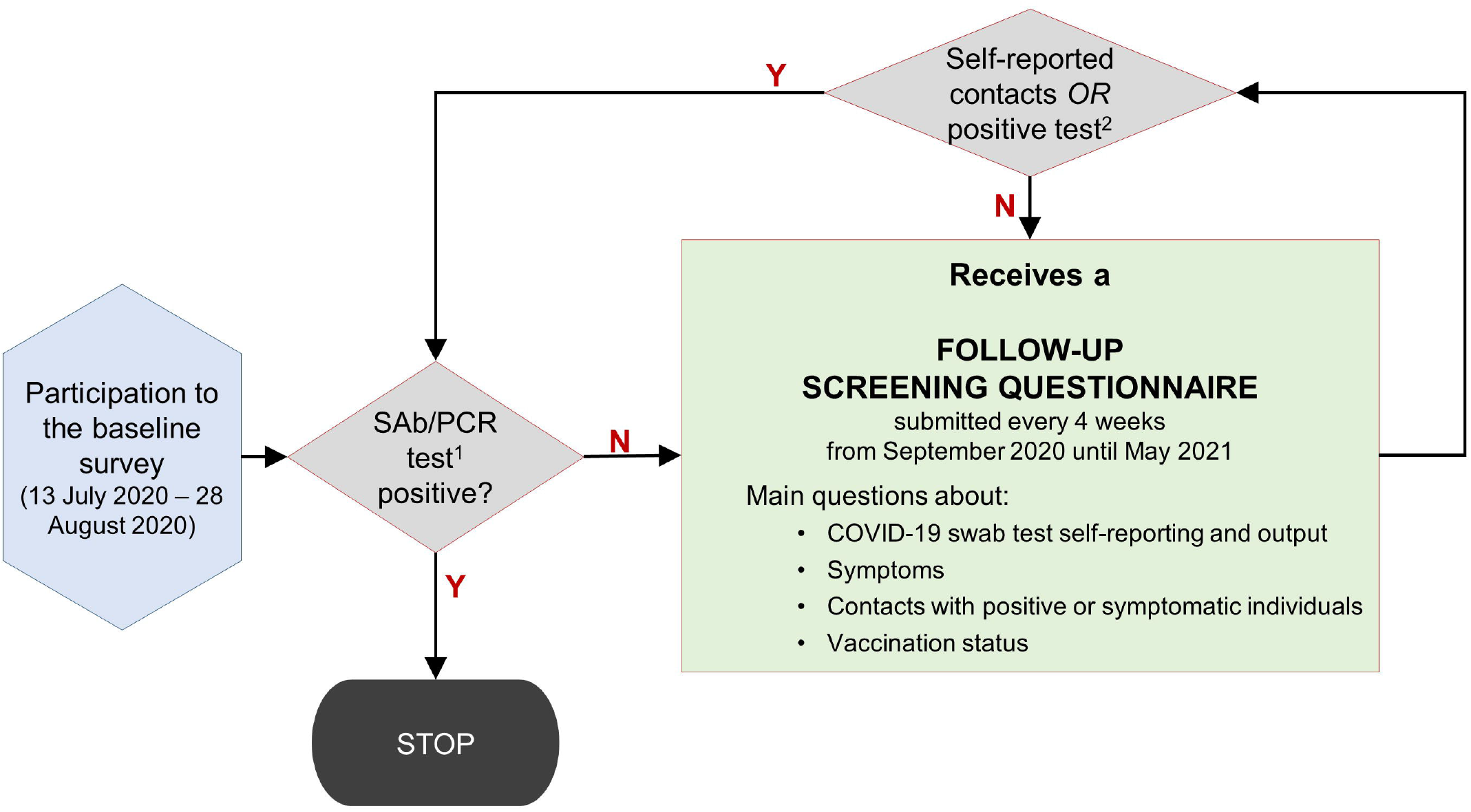
Flowchart of the follow-up screening in the CHRIS COVID-19 study from the individual participant point of view. Baseline participation involved 845 individuals. Of them, 9 were immediately excluded from the follow-up because of a positive test and 139 never replied to any follow-up questionnaire, leaving 697 individuals with available follow-up data who were included in the analyses. Detailed figures by month are reported in **Table 2**. ^1^Performed at the study centre. ^2^Self-reported contacts with positive or symptomatic individuals or any positive swab test.

At baseline, 845 individuals participated to the study, corresponding to 58.3% of the 1450 target sample size. Among participants, the youngest and the oldest age groups were less represented than in the non-respondents, and participants had a higher educational level (**Supplementary Table 1**). The participation bias was addressed through non-response sampling weights (see below), which enabled calibration to the Val Venosta/Vinschgau population age and sex distribution (**Supplementary Table 2**). Of the 845 baseline participants, 9 tested positive at baseline and 138 dropped out of the study by not replying to further questionnaires. One further individual and 21 questionnaire responses overall were also removed to prevent possible overlapping of a reported positive swab test in the questionnaire with the test performed at the study center. This left 697 individuals and 4,512 follow-up questionnaires available for analysis.

**Table 1.** Baseline characteristics of the 697 participants in the CHRIS Covid-19 study follow-up. Sample size (N) and relative frequency (%) or median (range) are given for categorical and quantitative variables, respectively.

| Characteristics | Values | N |
| --- | --- | --- |
| Age, years |  |  |
| Median (range) | 50 (19-93) | 697 |
| Sex, % (n=697) |  |  |
| Female | 48.6 | 339 |
| Male | 51.4 | 358 |
| Educational qualification, % (n=684) |  |  |
| Primary school or no title | 22.5 | 154 |
| Secondary school | 59.6 | 408 |
| University degree | 17.8 | 122 |
| Rooms in the house, No. |  |  |
| Median (range) | 3 (1-26) | 686 |
| Cohabitants, No. |  |  |
| Median (range) | 4 (1-6) | 689 |
| Main occupation, % (n=658) |  |  |
| Primary and secondary sector | 22.3 | 147 |
| Tertiary sector – Housekeeper | 6.5 | 43 |
| Tertiary sector – Education | 8.8 | 58 |
| Tertiary sector – Healthcare | 3.5 | 23 |
| Tertiary sector – Social services | 2.9 | 19 |
| Tertiary sector – Others | 30.5 | 201 |
| Student or unemployed | 3.0 | 20 |
| Retired | 22.3 | 147 |
| Smoking status, % (n=675) |  |  |
| Never smoker | 56.9 | 384 |
| Past smoker | 28.3 | 191 |
| Current smoker | 14.8 | 100 |
| Body mass index, $kg/m^2$ | | |
| Median (range) | 24.6 (17.7-52.2) | 693 |
| Any pathologies, % (n=676) |  |  |
| Yes | 49.9 | 337 |
| No | 50.2 | 339 |

**Table 2.**
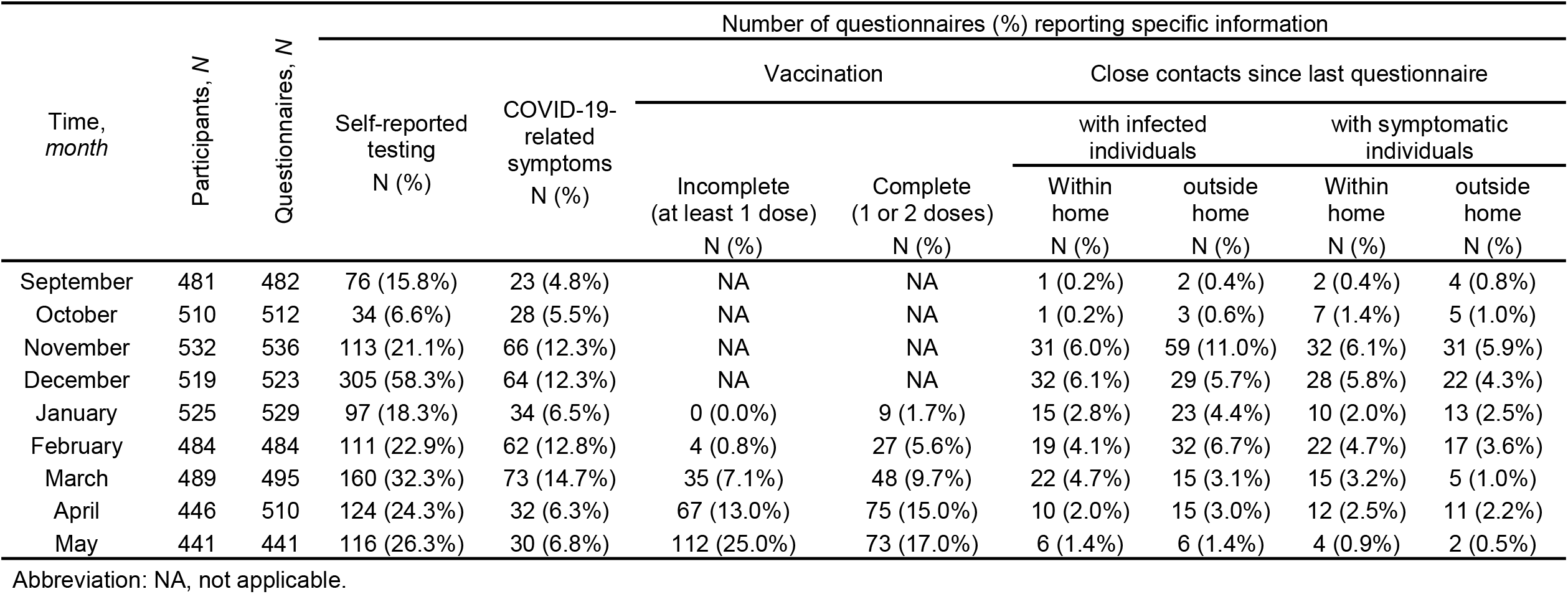
Distribution of study participants and questionnaires in the CHRIS COVID-19 study follow-up by month of participation, overall and by self-reported SARS-CoV-2 related information.

### Primary endpoint

Our primary interest was on the probability to have undergone a nasopharyngeal swab for SARS-CoV-2 testing, based on the question “*Have you had a naso/oropharyngeal swab for the novel coronavirus infection since the date of the last questionnaire?*” included in all follow-up questionnaires.

### Study variables

The probability of undergoing a swab test was assessed against time-invariant and time-varying characteristics collected during baseline and follow-up interviews, respectively. The groups of time-invariant characteristics were: a) biological characteristics: age, sex, and body mass index (BMI); b) any pre-existing pathologies (more details in the **Supplementary Methods**); c) lifestyle determinants: smoking status; and d) socio-demographic characteristics: educational qualification, main occupation, and crowding index (ratio between number of cohabitants and number of rooms in the house as a measure of household living conditions). The group of time-varying characteristics was: e) information reflecting individual symptomatology, close contacts with infected or symptomatic individuals defined as permanence within the same indoor environment for at least 15 minutes at a distance of less than 2 meters without protection or direct physical contact with another person, and SARS-CoV-2 vaccination status (23).

The month at follow-up questionnaire completion was used as a proxy for the general context, namely dynamic policies to address the spread of infection and the progression of the pandemic, among other unmeasured determinants. Details about all characteristics considered for the analyses are reported in the **Supplementary Methods** and elsewhere (23).

### Statistical analysis

Mixed-effect logistic regression models were fitted to investigate the association between the probability to undergo testing and several determinants, setting a random intercept on each participant. Models were fitted using the R package GLMMadaptive v0.8-5 (https://cran.r-project.org/package=GLMMadaptive) following the standard formulation 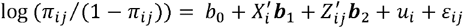, where *i* and *j* indicate the *i*^th^ individual at time *j*, π_*ij*_ indicates the probability of individual *i* to get tested at time *j, b*_0_ is an intercept term, *X*_*i*_ is a matrix formed by all time-invariant covariates (e.g.: sex, age, level of education, main occupation), *Z*_*ij*_ is a matrix formed by all time-varying covariates (e.g.: contacts with infected or symptomatic individuals), *u*_*i*_ is a normally distributed random effect on the individuals (random intercept), ε_*ij*_ is a normally distributed error term, and ***β***_1_ and ***β***_2_ are the vectors of fixed effect coefficients to be estimated (log odds ratios) for the time-invariant and time-varying covariates, respectively.

In the logistic regression models, to correct for potential selection bias, we included relative sampling weights based on the observed sex-age strata distribution in the study sample relative to the strata distribution in the target population (25), as outlined in the **Supplementary Methods**. We fitted models to separate the relative contribution of each block of determinants, which incrementally included: 1) the month of participation; 2) individual biological characteristics (age, sex, BMI) and pathologies; 3) lifestyle (smoking) and socio-demographic characteristics (educational qualification, main activity, and crowding index); 4) individual symptomatology; 5) contacts with infected or symptomatic individuals inside and outside home; 6) and vaccination status. Goodness of fit was assessed via the Akaike Information Criterion (AIC) and the proportion of variance explained by separate blocks was estimated by the marginal R-squared statistic (26). Marginalized coefficients and the corresponding standard errors were estimated using Monte Carlo iterations over the random effects with 1000 samples repeated 50 times (27). To assess the marginal effect of each determinant, we also fitted models adjusted for age, sex, and month at follow-up, where each determinant was included in turn. The full model was also stratified by early and late study periods (September-December 2020; January-May 2021) to account for the introduction of vaccinations on December 27^th^, 2020, and the potential changes in social and business restrictions(21).

Despite an apparent small amount of missing data overall (**Supplementary Figure 1**), the propagation of missing data from the baseline variables thorugh the longitudinal structure of the dataset implied substantial data loss: of 697 individuals with 4512 corresponding follow-up questionnaires, complete records were available only for 620 individuals (−11%), corresponding to 3826 follow-up questionnaires (−15%). The largest proportion of missing values was observed for ‘main occupation’, with 277 missing observations from 39 individuals, corresponding to 6.1% of the 4512 questionnaires. In 131 cases, ‘main occupation’ was the only variable with missing data. The joint distribution of missing data across all variables is depicted in **Supplementary Figure 2**. Assuming missingness at random (MAR), missing values were multiply imputed using chained equations (MICE) (28,29), based on 3 imputed datasets, exploiting the correlation between variables (**Supplementary Tables 3** and **4**). Extensive reasoning supporting MICE and respective methods are presented in the **Supplementary Methods**. Sensitivity analyses were conducted, increasing the number of imputations to 50 (**Supplementary Methods**). A complete case analysis (CCA) was also performed for completeness. All analyses were performed using the R software package version 4.0.5 (30).

## Results

Study participants had a median age of 50 years (range: 19, 93), 48.6% were females, 3.5% were healthcare workers, and 49.9% declared at least one pathology (**Table 1**). Between September 2020 and May 2021, the number of self-reported SARS-CoV-2 tests was 1,136 (median number of test per capita 1; range 0, 7). One-hundred-and-two individuals reported to have tested positive at least once, totaling 128 positive tests. The percentage of individuals self-reporting SARS-CoV-2 testing varied between 6.6% in October and 58.0% in December. The percentage of those reporting within-home contacts varied between 0.2% (September) and 6.1% (December), and between 0.4% (September) and 11.0% (November) for outside-home contacts (**Table 2**).

When fitting sequential models, the model that just included the month of participation had a marginal R-squared estimate 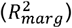 of 0.153 (AIC=4474.9), which increased to 0.169 (i.e.: +0.016; AIC=4452.7) when including individual biological characteristics (age, sex, and BMI) and pre-existing pathologies, and to 0.191 (additional +0.022; AIC=4441.3) when further ncluding lifestyle (smoking) and sociodemographic characteristics such as educational qualification, economic activity, and crowding index (**Supplementary Table 5**). When further adding the presence of symptoms (**Figure 2**; **Supplementary Table 5**), 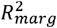 increased to 0.308 (+0.117 from previous model; AIC=3992.6) and to 0.387 when also adding information on close contacts (+0.079; AIC=3764.3). Final inclusion of vaccination status ((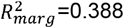 =0.388, AIC=3760.3) did not add substantial information (**Figure 2**; **Supplementary Table 5**). This last fully adjusted model suggested the following determinants to be associated with undergoing a SARS-CoV-2 test: close contacts with infected individuals within (OR:7.47, 95%CI:3.81–14.62) and outside home (OR:9.87, 95%CI:5.78–16.85), self-reported symptoms (OR:8.26, 95%CI:6.04–11.31), month of reporting (e.g. December vs. September OR:7.19, 95%CI:5.35– 9.66), main occupational activity (e.g.: retired individuals compared to individuals working in the ‘other’ category of the tertiary sector, OR:0.50 95%CI:0.34–0.73), and vaccination (OR:1.59, 95%CI:1.08–2.33 for individuals who initiated vaccination compared to non-vaccinated; **Figure 2**; **Supplementary Table 5**). For comparison, we provide results of a model adjusted only by month of participation, age, and sex, where each other variable was included singularly in **Supplementary Table 6**. Results were very similar to the full model, with additional significant associations observed for age, educational level, and contacts with symptomatic individuals.

**Figure 2.**
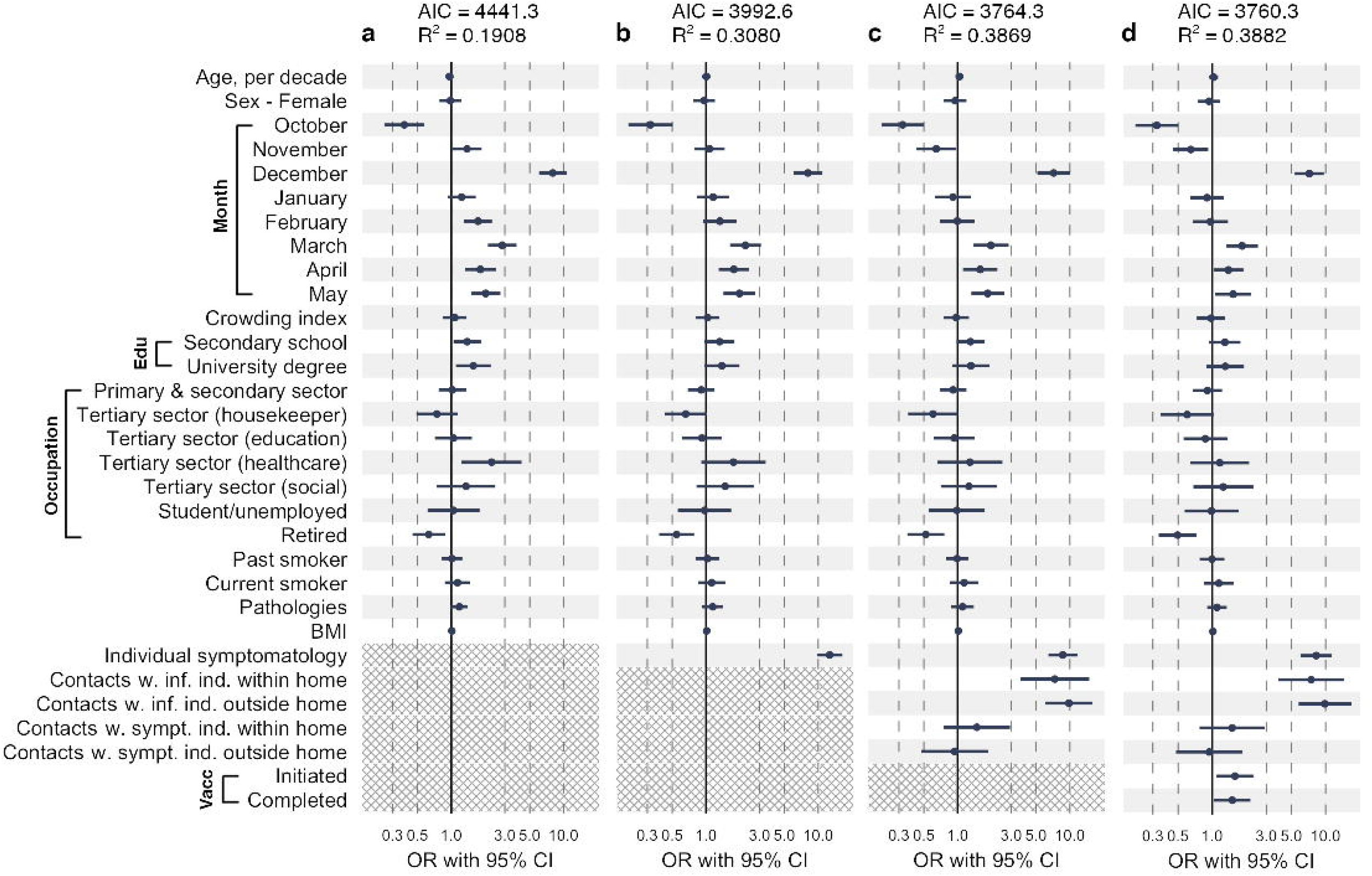
Results of the mixed-effect logistic regression models. Considered was the whole period from September 2020 until May 2021. The reference categories for the categorical variables were: ‘Male’ for Sex; ‘September’ for month; ‘Primary school or no title’ for Educational qualification; ‘Tertiary sector (other)’ for main occupation; ‘Never smoker’ for smoking status; and ‘Not vaccinated’ for vaccination status. Abbreviations: OR, odds ratio; CI, confidence interval. Shading is used to highlight variables not included. On top of each panel the pseudo R^2^ and the AIC statistics are reported (see Methods). **Panel a**: Included are all time-invariant variables collected at baseline and month of participation. **Panel b**: In addition to variables included in previous panel, individual symptomatology is included. **Panel c**: Variables indicating contacts with infected and with symptomatic individuals are further included. **Panel d**: Additional inclusion of vaccination information.

Age and educational level were correlated and they were also correlated with other variables such as occupation. Contacts with symptomatic individuals was strongly correlated with contacts with positive individuals, therefore they were associated with the testing probability singularly but not independently when both were included in the full model. In the age-, sex- and month-adjusted model, we also observed that the ORs of testing given contacts with positive individuals were larger than the ORs of testing given contacts with symptomatic individuals, meaning that knowing that a contact was positive was a stronger determinant of testing compared to knowing that he/she was symptomatic. We also observed a positive association with the month of November. This association was similar to the model adjusted for time-invariant variables (**Figure 2A**). Then, the effect of November became null when further adjusting for symptomatology (**Figure 2B**) and reversed when further including contacts with infected or symptomatic individuals (**Figure 2C** and **2D**). This indicates that those who were tested were also those presenting symptoms, who were those who had contacts with infected or symptomatic individuals.

Results of the full model were unchanged when running a MICE based on 50 rather than 3 imputations (**Supplementary Table 7**). Conversely, a CCA would have implicated somewhat different results for variables with most missing data such as ‘main occupation’, underlining that the hypothesized missingness model may influence the conclusions (**Supplementary Table 8**).

To reflect the vaccination campaign that started on December 27, 2020, the full model was stratified by period: from September to December 2020 and from January to May 2021. Results were consistent with those of the overall analyses (**Supplementary Table 9**). In the first period, the determinant most associated with SARS-CoV-2 testing was within-home close contacts with infected individuals (OR:9.98, 95%CI:3.81–26.13). In the latter period, this changed to having outside-home close contacts with infected individuals (OR:14.65, 95%CI:6.71–32.00), while within-home close contacts with symptomatic individuals was still associated, but to a minor extent (OR:3.98, 95%CI:1.37–11.57). In the second period, vaccination was associated with SARS-CoV-2 testing (OR:1.81 with 95%CI:1.25–2.62 for initiated vaccination versus no vaccination; OR:1.62, 95%CI:1.08–2.45 for completed vaccination versus no vaccination; **Supplementary Table 9**) more clearly than in the overall model.

## Discussion

This longitudinal analysis of a population-representative sample showed that the probability to undergo SARS-CoV-2 testing was mostly driven by contextual rather than individual characteristics. The major determinants were the time of the year, which reflects a mixture of the pandemic trend and public health interventions like restrictions or more intense testing activities, and close contacts with infected and symptomatics individuals. Within-home close contacts were most strongly associated with SARS-CoV-2 testing until December 2020, while outside-home close contacts were most strongly associated with testing probability from January 2021 onward, probably reflecting a change in the pandemic mitigation policies between the two periods. Among the broad spectrum of individual determinants considered here, only retirees showed evidence of reduced testing probability as compared to other main occupational groups. No established personal characteristic among those previously associated with severe COVID-19 disease, such as age, BMI, smoking status and pre-existing pathologies, were associated with SARS-CoV-2 testing.

An important feature of the study was the population-representative sampling, which enables the generalization of the findings to the whole district and perhaps to the wider rural areas of South Tyrol. Caution is recommended for extrapolating the results to other contexts: the characteristics of the rural Alpine district considered here (24), which has relatively low population density and strong administrative autonomy, can be substantially different from those of urban environments or culturally diverse regions.

Of note is that the study was conducted in a COVID-19 near-naïve population: until the beginning of our assessment, the district had as few as 16 SARS-CoV-2 confirmed cases (31). This contingency allowed to observe participants prospectively on a monthly basis to assess incident cases of first-time infection. Upon confirmation of a SARS-CoV-2 infection, among participants prospectively recalled at the study center based on their risk profile by questionnaire self-report, participants would exit the follow-up screening and enter a separate assessment protocol, which is beyond the current scope and fully described elsewhere (23). On the one hand, this design could have limited the breadth of participants undergoing and reporting further testing for possible re-infection or confirmation/denial of prior test/exposure. On the other hand, it has allowed to focus the selection criteria on a relatively homogeneous population susceptible to first-time infection.

A major strength of the study was the regular follow-up conducted over nearly a whole year of the COVID-19 pandemic. The frequency of follow-up questionnaires, submitted every four weeks, should have largely limited recall bias and enhanced data quality.

In addition to the limited sample size, which prevented the possibility to study higher order interactions, other limitations should be highlighted. First, most information was self-reported, thus some reporting error could not be excluded. A subtle implication of the self-reported information is that the study sample may have not only included susceptible individuals. Individuals who did not test or who did not report a positive test or a contact with a known or unknown positive individual might have still been included without any assessment of their infectious status. On the other hand, an individual being possibly unaware of own prior infection status it might not alter attitude or predisposition to testing, in the absence of symptoms or contacts with infected persons, as investigated in our study. Second, even if individuals were randomly selected by stratification and the analyses accounted for population-representative weights, some selection bias cannot be excluded, although we counteracted it by applying relative sample weights stratified by the age and sex distribution of the target population (4,23). Third, we did not consider availability of and distance to testing sites, which may affect the chance to undergo a test (32). Distance was only available as an average distance at the municipal level, and testing sites could also have changed overtime, depending on the intensity of the pandemic. Fourth, our study was not designed to incorporate external events such as public health interventions or hospital admissions, which could only be approximated by modeling the month of participation. Furthermore, ‘month’ was a proxy for other unmeasured factors, which are all likely to impact the probability of testing, including the transmission intensity, testing recommendations, and testing availability. Fifth, the widespread availability of cheap, rapid, nasal tests through pharmacies, without the need of a medical prescription, made it unfeasible to evaluate the role of general practitioners or other healthcare providers in mediating access to swab tests. Lastly, individuals who tested more frequently could have been more likely to test positive and thus being excluded from the study, introducing a selection bias. However, focusing on testing attitudes in our work was favored by a homogeneous selection of first-time infection susceptible participants. In fact, survivors that may have been confirmed positive in the past, may be partly protected from re-infection or severe disease by their immune response, especially in the short term. Purportedly, these circumstances may determine a different attitude towards further testing even in the event of contacts with infected persons or suffering from any symptoms. Prior positive or tracked participants may even be subject to and report testing to exit isolation or quarantine periods, which would exclude a major role for their attitude to testing. We have also included vaccination as possible intervening pathway, as a proxy for health-prone behavior before any confirmed infection, independent of possible indication of being prioritized for vaccination for professional or health-related reasons.

We identified December as the month with the highest reporting of SARS-CoV-2 testing, likely due to the mass testing implemented in South Tyrol at the end of November 2020 (22). The month in which the questionnaire was filled, social contacts with infected individuals and symptoms played a central role in undergoing SARS-CoV-2 testing. These findings support that individuals largely followed the general recommendations for undertaking a test when they had symptoms or contact with infected individuals, as suggested by health authorities, following testing prioritization in case of limited resources outlined by the WHO and ECDC (33–35).

The analyses stratified by period, before and after the start of the vaccination campaign, showed consistent results with the overall analysis, indicating a general stability of the identified determinants over the pandemic course. The main determinants associated with SARS-CoV-2 testing were within-home close contacts with infected individuals in the first period and outside-home close contacts with infected individuals in the second period. The effect of tightened measures to mitigate the spread of infection might have confined secondary infections within households from September to December 2020. For instance, in November 2020, in many Italian municipalities, including several municipalities in the study area considered here, a “red zone” policy was establish to minimize social interactions and drop the contagion rates (21). From January to May 2021, social measures were relaxed: higher order schools and non-essential businesses reopened, and mobility across Italian regions was gradually possible, therefore contacts with infected individuals outside home were more common (21). In this regard, we observed the reversing effect of November due to adjustment for symptomatology and contacts, indicating high correlation between this month, the presence of symptoms and contacts with infected or symptomatic individuals. Why such an overlapping situation was observed in November and not in the other months is also probably due to changes in the containment policies: until October 2020, containment measures were relaxed following the apparently controlled situation of low viral circulation observed until the summer 2020. This had favored interpersonal contacts. Furthermore, until the end of October 2020, rapid antigen tests were not available, and testing was limited to symptomatic individuals or those who got in contact with suspicious cases. In the following months, with a more widespread availability of rapid tests, the strict link between exposure to contacts – symptoms – testing might have been decoupled, with more people testing independently of symptoms (for instance, negative tests were required to go to the workplace or to attend events).

Also vaccination was associated with a higher probability to undergo SARS-CoV-2 testing. The elderly, the fragile, the healthcare workers, and the educational employees were the first categories invited for vaccination. These groups were also those with had a higher probability of being tested in the analyses adjusted only for age, sex and month. The increased risk of testing associated with vaccination might look surprising. To try to explain it, we should consider that study participants were reporting what had happened in the last 4 weeks and that the study was mainly carried out before large-scale vaccine administration in Italy, with steep increase of vaccination rates since spring of 2021, close to the end of our observation period. A first possibility is that, at these initial stages, those who were more prone to get vaccinated were also those who had a higher attitude to follow the health authority recommendations: they might have been taking a test prior to the vaccination, implicating the positive association between the two variables. However, during the first months of 2021, specific population groups were prioritized to get the vaccine (namely healthcare professionals, vulnerable individuals, the elderly). Thus, despite our analyses adjusted for age, chronic conditions, BMI and other socio-economic determinants, this explanation might not hold fully. A more likely explanation is that a negative test was required or highly recommended to prevent previously or extant infected individuals from attending vaccination, which would make the observed direction of the association possible.

The only personal characteristic associated with testing probability was being retired, which had lower odds of getting tested compared to all other occupational categories. The association was significant in the model adjusted only for age, sex, and month, but also in fully adjusted model with an even larger effect. Thus, this association was independent of all other variables included in the model. This effect might have been further enhanced by the retired group lacking individuals living in long-term care facilities (LTCF). While living in a LCTF was associated with very high odds of COVID-19 diagnosis and hospitalization (36), the CHRIS COVID-19 study experienced difficulty to enroll individuals residing in LTCFs due to lack of individual autonomy and legal responsibilities. It is thus likely that the retired group in the CHRIS COVID-19 study was composed of individuals in good health, living in their own houses with limited social interactions. A “green pass effect”, where unvaccinated workers had to undergo repeated, negative tests to get to the workplace, while retirees were perhaps less affected by this provision, can also be excluded: in Italy, the green pass came into force by October 15, 2021, that is, after the end of the study.

Perhaps surprising is the finding that healthcare workers did not have higher testing probability, when other studies suggested that healthcare workers should be more likely to undergo SARS-CoV-2 testing because they are more likely to have at-risk contacts and are at higher risk to infect vulnerable people (10,37,38). However, when limiting adjustment to age, sex, and month, without accounting for being in regular contact with infected or symptomatic individuals like patients in healthcare facilities, this effect was apparent.

In conclusion, among individuals susceptible to first SARS-CoV-2 infection, almost no personal characteristic, not even educational level or pre-existing pathologies, had an infuence on the likelihood of undergoing SARS-CoV-2 testing in the Val Venosta/Vinschgau district over nine months of the COVID-19 pandemic. An exception was being retired, which was related to lower testing probability. Was this type of knowledge from the general community available along with test results and personal risk assessments in a rigorous and timely fashion, it could inform testing prioritization and communication strategies (39,40). The homogeneous distribution of testing across different citizen categories may question public decision-makers as to whether the most at-risk groups were correctly prioritized during the most intense phases of the pandemic (41,42).

## Data Availability

All data produced in the present study are available upon reasonable request to the authors.

## Acknowledgements

We are grateful to all CHRIS COVID-19 study participants. We thank the collaborators of both Eurac Research and the Healthcare System of the Autonomous Province of Bolzano for field and laboratory operations, the representatives and staff of the local Administrative Authorities for support and all ancillary volunteers who eventually made the study possible.

## Data availability

Given national legal restrictions for the publication of health data, the Institute for Biomedicine developed an access policy compliant to it. The data used for this work can be obtained by submitting a substantiated request to the CHRIS Access Committee at.

## Authors’ contributions

CP, PPP, RM, and MG conceived the CHRIS COVID-19 study.

CP and DG conceived the investigation topic object of the present work.

DG, GB, MG, and DB performed the statistical analyses.

DG, GB, MG, DB, LF, CXW, RM and CP interpreted the results.

DG, RM, and CP drafted the manuscript.

LF, LB, RL, DB, RM, MG, CXW, PP contributed to the critical revision and editing of the manuscript.

