## Supplementary material for "Determinants of SARS-CoV-2 nasopharyngeal testing in a rural community sample susceptible of first infection: the CHRIS COVID-19 study"

### **Supplementary Methods**

#### **Determinants included in the analysis**

The questionnaires from which questions were extracted are fully available as a supplement of reference (1).

##### *Smoking status*

Information about smoking status was retrieved from the question 7.1 of the baseline questionnaires of the CHRIS COVID-19 study (1): *“Do you smoke cigarettes (or tobacco) in general?”*. The possible answers were: “I now smoke (at least once a day)”; “I occasionally smoke (less than once a day)”; “I smoked in the past and have now quit altogether”; “I have never smoked (less than 100 cigarettes in a lifetime)”; and “Prefer not to respond/Do not know”. In the analyses, we defined 3 smoking status levels:

- “No smoker”, including individuals answering “I have never smoked (less than 100 cigarettes in a lifetime)”;
- “Current smoker”, including individuals answering “I now smoke (at least once a day)”;
- “I occasionally smoke (less than once a day)”; and “Past smoker”, including “I smoked in the past and have now quit altogether”. If individuals reported to have smoked in the past, information about when they stop smoking was available the question 7.1.2 *“How long ago did you stop smoking?”*. We assumed “Past smoker” individuals stopping smoking less than a month before the interview, otherwise they were set to “Current smoker”;
- Responses “Prefer not to respond/Do not know” were set to missing.

##### *Educational qualification*

Educational qualification information was retrieved from the baseline questionnaire. Individuals could answer: “No title”; “Primary school”; “Lower secondary school”; “Vocational school”; “Certificate of upper secondary school”; “Bachelor’s degree or equivalent”; “Master or higher degree”; and “Prefer not to respond/Do not know”. We defined the following educational levels:

- “Primary school or no title” including individuals answering “No title” or “Primary school”;
- “Secondary school” including “Lower secondary school”, “Vocational school”, and “Certificate of upper secondary school”;
- “University degree” including “Bachelor’s degree or equivalent” and “Master or higher degree”.
- “Prefer not to respond/Do not know” were set to missing.

##### *Main sector of activity*

Main sector of activity information was retrieved from the questions 6.1 (“*In the 12 months before the coronavirus-related emergency, what was your primary occupational status?*”) and 6.2 (*“In the 12 months before the coronavirus-related emergency, what was your main sector of activity?*”) in the baseline questionnaire (1).

The possible closed answers to the 6.1 question were: “Stable occupation (including working for a family business or temporarily absent/on leave from work)”; “Temporary or seasonal occupation”; “Unemployed seeking occupation”; “Student”; “Retired”; “Housekeeper”; “Other situation”; and “Prefer not to respond/Do not know”.

The possible closed answers to the 6.2 question were: “Agriculture, forestry and farming”; “Mining, construction and industry”; “Craftsmanship”; “Provision of essential services, Transport and delivery”; “Accommodation and catering”; “Trade”; “Healthcare”; “Social services”; “Education”; “Public administration”; “Police, armed and public security force”; Other sector”; and “Prefer not to respond/Do not know”.

We considered the answers to the question 6.2 irrespective whether individuals answered to have a “Stable” or “Temporary or seasonal” occupation to the question 6.1 in order to define the main sectors of activity in the analyses. Therefore, we defined the following main sector of activities:

- “Primary and secondary sector”, including “Agriculture, forestry and farming”, “Mining, construction and industry”, and “Craftsmanship”;
- “Tertiary sector – healthcare”, including “Healthcare”;
- “Tertiary sector – education”, including “Education”;
- “Tertiary sector – social services”, including “Social services”;
- “Tertiary sector – others”, including “Provision of essential services, Transport and delivery”, “Accommodation and catering”, “Trade”, “Police, armed and public security force”, and “Other sector”.
- Housekeeper individuals (“Housekeeper”) were defined using “Housekeepers” answers to question 6.1 among those answering “Prefer not to respond/Do not know” to question 6.2.
- Students and unemployed individuals (defined as “Student or unemployed” in the analyses) were identified using “Unemployed seeking occupation” or “Student” to question 6.1 among those answering “Prefer not to respond/Do not know” to question 6.2.
- The remaining individuals reported “Prefer not to respond/Do not know” in either question 6.1 or 6.2 were set to missing.

##### *Close contact with infected and/or with symptomatic individuals*

Information about close contact with infected and with symptomatic individuals was retrieved from the following questions in the baseline and follow-up questionnaires: *“Have you been in close contact with someone with coronavirus infection living with you?”;* *“Have you been in close contact with someone with coronavirus infection NOT living with you?”*; *“Have you been in close contact with someone with symptoms living with you?”*; and “*Have you been in close contact with someone with symptoms NOT living with you?”.* A note next to the questions explained that ‘close contact’ had to be interpreted as permanence within the same indoor environment for at least 15 minutes at a distance of less than 2 meters without protection otherwise physical contact with another person.

##### *Symptomatology*

Individual symptomatology information was retrieved from the following question present in the follow-up questionnaires: *“Except for possible symptoms you may regularly suffer from, have you had any of the following symptoms, since the last questionnaire?”*(1). The 26 listed symptoms were: fever; shivers or chills; fatigue or tiredness; joint or muscle pain; headache; lack of appetite; loss of taste; loss of smell; ear pain (otitis); redness or burning eyes; eye pain; cold; sore throat or hoarseness; dry cough; wet cough; coughing up blood; shortness of breath; chest pain; tachycardia or palpitations; abdominal pain; nausea; vomiting; diarrhea; pale or oily feces; skin hypersensitivity; and itching or rash.

##### *Pathologies*

Information about pre-existing pathologies was retrieved from the following question in the baseline questionnaire: *“Has a doctor ever diagnosed you with any of the following diseases?”*(1). The listed pathologies were: diabetes; other metabolic dysfunction; respiratory allergy; other type of allergy; asthma; chronic bronchitis; other lung disease (excluding cancer); hypertension; arrhythmia; ischemic or cerebrovascular disease; other cardiovascular disease; kidney disease; liver disease; autoimmune rheumatic disease; musculoskeletal disease; blood disease; mental or affective disease; and cancer.

#### **Derivation of relative sampling weights**

The relative sampling weights included in the logistic regression models were derived as follows:

- Let set i = 1, 2 as an index for sex (1 = males; 2 = females) and j = 1..6 as an index for the age categories (18-34; 35-44; 45-54; 55-64; 65-74; 75+ years).
- At the population level, the relative frequency of a specific stratum was defined as RF_ij_ = AF_ij_ / N, where N was the population size and AF_ij_ the absolute frequency of the stratum (i,j) in the population.
- At the study sample level, the relative frequency of a specific stratum was defined as rf_ij_ = af_ij_ / n, where n was the sample size and af_ij_ the absolute frequency of the stratum (i,j) in the study sample.
- The relative sample weights (rsw) were then estimated as rsw_ij_ = RF_ij_ / rf_ij_.

#### **Multiple imputation of missing values**

The percentage of missing values across predictors varied between 0.4% (symptomatology) and 5.6% (main sector of activity). In longitudinal studies, missing data may be present for baseline, time-invariant predictors such as sex, age, level of education, or occupation (defined as level-1 predictors) and for longitudinal, time-varying predictors collected during the study course such as symptomatology, contacts with infected and symptomatic individuals or vaccination status (level-2 predictors) (2,3). In our study, only few variables presented missing data. However, the number of missing observations that would have been excluded from the analyses could have been due to missing values propagation from missing baseline characteristics such as Main Occupation and Level of Education through the longitudinal structure of the study. An analysis limited to records with no missing data (complete case analysis, CCA), would have included 620 individuals with 3,826 follow-up questionnaires. In contrast, the multiple imputation (MI)-based analysis enabled exploitation of data from all 697 individuals with 4,512 corresponding questionnaires. This difference corresponded to a loss of 11% individuals and 15% questionnaires in the CCA compared to the MI analysis.

The marginal pattern of missing data is illustrated in **Supplementary Figure 1**. The largest proportion of missing values was observed for Main Occupation, with 277 missing observations from 39 individuals, corresponding to 6.1% of the 4,512 questionnaires. The upset plot in **Supplementary Figure 2** shows that, out of these 277 cases, for 131 of them Main Occupation was the only variable affected. Contacts with infected and/or symptomatic individuals at home were the next variables, in terms of percent of missing values. This means that a CCA could exclude a large number of records due to missing values from a single variable.

We found it reasonable to assume that data could generally be missing at random (MAR) rather than missing completely at random (MCAR). Under MAR, if there were missing data following MCAR mechanisms, accuracy of estimates would not be affected as the covariate in the imputation model would have near-zero correlations with the missing values.

Supplementary Tables 3 and 4 show the distribution of Main Occupation by Sex and Level of education, respectively. The distribution of Main Occupation was associated with Sex. For example, women were more likely to work as housekeepers and in the fields of education and healthcare. Men were more likely to work in the primary or secondary sector (Supplementary Table 3). For this reason, sex was included as a predictor in the imputation model to impute missing data of Main Occupation. The same reasons were valid for the educational level. The probability to work in the Tertiary Sector (Education) was higher among individuals with a university degree (36%) compared to individuals with primary or secondary level of education (0% and 5.7%), respectively (Supplementary Table 4). Individuals with primary level of education were more likely to be retired (48%) than those with higher level of education: retirees were 17.3% among those with a secondary level of education and 7.6% among those with university degree (Supplementary Table 4).

The option to recode missing values of categorical variables as a separate category, to limit the sample size loss, was discarded given extensive literature highlighting that this approach would very likely bias the results (4,5).

We used three imputed datasets based on multiple imputation chained equations (MICE) using 50 iterations. This technique improves accuracy and statistical power under MAR assumption. The imputation order of variables proceeded in ascending order of the proportion of missing values. In the imputation procedure, we also incorporated auxiliary information as past serological tests and extant pathologies. Continuous, binary, and categorical variables were imputed using two-level predictive mean matching, binary and polytomous logistic regression, respectively. Month was defined as a cluster variable. Every target variable with missing values was imputed including different candidate predictors using an automated procedure based on the correlation structure underlying the data. For instance, Main Occupation was multiply imputed including Sex and Level of education as predictors in the imputation model. For each variable pair, the procedure calculated two correlations using pairwise complete observations. The first one was the correlation between the imputation target variable and the candidate predictor variable. The second one was the correlation between the missing indicator of the target variable (0=observed;1=missing) and the candidate predictor variable. If the largest absolute value of these double pairwise correlations between a target variable and a candidate predictor exceeded 0.10, the predictor was included in the imputation set for the target variable with missing values. This procedure was performed using the function *quickpred* in the R package ‘mice’ version 3.13.0 (used for the imputation procedure) using the default threshold of |0.10|. The crowding index variable was calculated after imputation of the number of cohabitants and the number of rooms (post-processing imputation). Imputation was performed with the R package ‘mice’ version 3.13.0, which used Rubin’s rules (6) to combine the estimates.

**Supplementary Table 1.** Distribution of the 1812 randomly selected candidates by sex, age group, and educational qualification, according to response status in the CHRIS COVID-19 study and overall.

| **Stratification variable** | **Strata** | **Non-participants** | | **Participants** | | **Total** | |
| --- | --- | --- | --- | --- | --- | --- | --- |
|  |  | **No.** | **%** | **No.** | **%** | **No.** | **%** |
| Sex; Age-group (years) | M; 18-34 | 155 | 16.03% | 95 | 11.24% | 250 | 13.80% |
|  | M; 35-44 | 78 | 8.07% | 72 | 8.52% | 150 | 8.28% |
|  | M; 45-54 | 83 | 8.58% | 94 | 11.12% | 177 | 9.77% |
|  | M; 55-64 | 82 | 8.48% | 68 | 8.05% | 150 | 8.28% |
|  | M; 65-74 | 37 | 3.83% | 64 | 7.57% | 101 | 5.57% |
|  | M; 75+ | 51 | 5.27% | 28 | 3.31% | 79 | 4.36% |
|  | F; 18-34 | 138 | 14.27% | 88 | 10.41% | 226 | 12.47% |
|  | F; 35-44 | 67 | 6.93% | 75 | 8.88% | 142 | 7.84% |
|  | F; 45-54 | 94 | 9.72% | 87 | 10.30% | 181 | 9.99% |
|  | F; 55-64 | 64 | 6.62% | 77 | 9.11% | 141 | 7.78% |
|  | F; 65-74 | 43 | 4.45% | 52 | 6.15% | 95 | 5.24% |
|  | F; 75+ | 75 | 7.76% | 45 | 5.33% | 120 | 6.62% |
|  |  | 967 | 100% | 845 | 100% | 1812 | 100% |
| Education* | up to lower secondary education | 269 | 27.85% | 195 | 23.08% | 464 | 25.62% |
|  | professional qualification | 412 | 42.65% | 328 | 38.82% | 740 | 40.86% |
|  | upper secondary education | 223 | 23.08% | 212 | 25.09% | 435 | 24.02% |
|  | university degree | 62 | 6.42% | 110 | 13.02% | 172 | 9.50% |
|  |  | 967 | 100% | 845 | 100% | 1812 | 100% |

*Based on information available in the CHRIS study prior to sample the participants

**Supplementary Table 2.** Raw and weighted distribution of the 845 baseline participants in the CHRIS COVID-19 study across sex and age group selected by design compared to the reference population.

| **Sex; Age-group (years)** | **CHRIS COVID-19** | | |  | **Population** | |
| --- | --- | --- | --- | --- | --- | --- |
|  | **No.** | **Raw %** | **Weighted %** |  | **No.** | **%** |
| M; 18-34 | 95 | 11.24% | 13.56% |  | 3,965 | 13.56% |
| M; 35-44 | 72 | 8.52% | 8.08% |  | 2,364 | 8.08% |
| M; 45-54 | 94 | 11.12% | 9.78% |  | 2,861 | 9.78% |
| M; 55-64 | 68 | 8.05% | 8.32% |  | 2,433 | 8.32% |
| M; 65-74 | 64 | 7.57% | 5.44% |  | 1,591 | 5.44% |
| M; 75+ | 28 | 3.31% | 4.87% |  | 1,425 | 4.87% |
| F; 18-34 | 88 | 10.41% | 12.44% |  | 3,637 | 12.44% |
| F; 35-44 | 75 | 8.88% | 7.62% |  | 2,227 | 7.62% |
| F; 45-54 | 87 | 10.30% | 9.60% |  | 2,807 | 9.60% |
| F; 55-64 | 77 | 9.11% | 8.08% |  | 2,362 | 8.08% |
| F; 65-74 | 52 | 6.15% | 5.22% |  | 1,525 | 5.22% |
| F; 75+ | 45 | 5.33% | 6.99% |  | 2,043 | 6.99% |
|  | 845 | 100% | 100% |  | 29,240 | 100% |

**Supplementary Table 3.** Distribution of Main Occupation by Sex.

| **Main Occupation** | **Males (n = 339)** | | **Females (n = 358)** | |
| --- | --- | --- | --- | --- |
|  | **n** | **%** | **n** | **%** |
| Primary and secondary sector | 121 | 35.7 | 26 | 7.3 |
| Tertiary sector (housekeeper) | 3 | 0.9 | 40 | 11.2 |
| Tertiary sector (education) | 10 | 2.9 | 48 | 13.4 |
| Tertiary sector (healthcare) | 5 | 1.5 | 18 | 5.0 |
| Tertiary sector (social) | 2 | 0.6 | 17 | 4.7 |
| Tertiary sector (others) | 97 | 28.6 | 104 | 29.1 |
| Student or unemployed | 8 | 2.4 | 12 | 3.4 |
| Retired | 74 | 21.8 | 73 | 20.4 |
| Missing | 19 | 5.6 | 20 | 5.6 |

**Supplementary Table 4.** Distribution of Main Occupation by Level of Education.

| **Main Occupation** | **Primary school or no title (n = 154)** | | **Secondary school**  **(n = 408)** | | **University degree**  **(n = 122)** | |
| --- | --- | --- | --- | --- | --- | --- |
|  | **n** | **%** | **n** | **%** | **n** | **%** |
| Primary and secondary sector | 32 | 21.8 | 95 | 23.3 | 19 | 15.6 |
| Tertiary sector (housekeeper) | 10 | 6.5 | 26 | 6.4 | 7 | 5.7 |
| Tertiary sector (education) | 0 | 0.0 | 22 | 5.4 | 36 | 29.5 |
| Tertiary sector (healthcare) | 1 | 0.6 | 15 | 3.7 | 7 | 5.7 |
| Tertiary sector (social) | 1 | 0.6 | 13 | 3.2 | 5 | 4.1 |
| Tertiary sector (others) | 33 | 21.4 | 142 | 34.8 | 22 | 18.0 |
| Student or unemployed | 0 | 0.0 | 7 | 1.7 | 13 | 10.7 |
| Retired | 71 | 46.1 | 67 | 16.4 | 9 | 7.4 |
| Missing | 6 | 3.9 | 21 | 5.1 | 4 | 3.3 |

**Supplementary Table 5.** Results from the block adjusted logistic mixed-regression models. Model 1 included only month. Model 2 additionally included individual biological characteristics (age, sex, and BMI) and pathologies; Model 3 additionally included lifestyle (smoking) and socio-demographic characteristics (educational qualification, main activity, and crowding index). Model 4 additionally included individual symptomatology. Model 5 additionally included information about close contacts with infected and symptomatic individuals within and outside home. Model 6 additionally included vaccination status.

| **Variable** | | | **Model 1** | | | **Model 2** | | | **Model 3** | | | **Model 4** | | | **Model 5** | | | **Model 6** | | |
| --- | --- | --- | --- | --- | --- | --- | --- | --- | --- | --- | --- | --- | --- | --- | --- | --- | --- | --- | --- | --- |
|  |  |  | **AIC** = 4474.9  **R**^2^_marg_ = 0.153 | | | **AIC** = 4452.7  **R**^2^_marg_ = 0.169 | | | **AIC** = 4441.3  **R**^2^_marg_ = 0.191 | | | **AIC** = 3992.6  **R**^2^_marg_ = 0.308 | | | **AIC** = 3764.3  **R**^2^_marg_ = 0.387 | | | **AIC** = 3760.3  **R**^2^_marg_ = 0.388 | | |
|  |  |  | **OR** | **L95** | **U95** | **OR** | **L95** | **U95** | **OR** | **L95** | **U95** | **OR** | **L95** | **U95** | **OR** | **L95** | **U95** | **OR** | **L95** | **U95** |
| Month  (ref.  September) | | October | 0.39 | 0.26 | 0.60 | 0.39 | 0.26 | 0.58 | 0.38 | 0.25 | 0.57 | 0.32 | 0.20 | 0.50 | 0.32 | 0.21 | 0.50 | 0.32 | 0.21 | 0.50 |
|  |  | November | 1.38 | 1.00 | 1.90 | 1.38 | 1.02 | 1.86 | 1.38 | 1.02 | 1.87 | 1.07 | 0.79 | 1.47 | 0.65 | 0.43 | 0.97 | 0.65 | 0.45 | 0.92 |
|  |  | December | 7.59 | 5.99 | 9.62 | 7.82 | 5.96 | 10.27 | 8.04 | 6.04 | 10.70 | 8.10 | 6.01 | 10.91 | 7.17 | 5.11 | 10.07 | 7.19 | 5.35 | 9.66 |
|  |  | January | 1.24 | 0.90 | 1.71 | 1.24 | 0.91 | 1.68 | 1.24 | 0.92 | 1.66 | 1.16 | 0.83 | 1.62 | 0.91 | 0.63 | 1.31 | 0.90 | 0.64 | 1.26 |
|  |  | February | 1.72 | 1.26 | 2.35 | 1.72 | 1.29 | 2.30 | 1.73 | 1.30 | 2.30 | 1.33 | 0.95 | 1.87 | 0.99 | 0.70 | 1.42 | 0.97 | 0.68 | 1.38 |
|  |  | March | 2.71 | 2.12 | 3.46 | 2.80 | 2.07 | 3.78 | 2.85 | 2.12 | 3.81 | 2.24 | 1.63 | 3.08 | 1.98 | 1.37 | 2.85 | 1.84 | 1.36 | 2.58 |
|  |  | April | 1.78 | 1.33 | 2.38 | 1.80 | 1.32 | 2.45 | 1.82 | 1.32 | 2.50 | 1.77 | 1.30 | 2.41 | 1.59 | 1.12 | 2.26 | 1.39 | 1.08 | 1.97 |
|  |  | May | 1.97 | 1.49 | 2.59 | 2.00 | 1.44 | 2.77 | 2.03 | 1.50 | 2.74 | 1.98 | 1.43 | 2.75 | 1.85 | 1.33 | 2.59 | 1.53 | 1.06 | 2.20 |
| Age, per decade | | |  |  |  | 0.85 | 0.80 | 0.90 | 0.96 | 0.88 | 1.05 | 1.01 | 0.92 | 1.10 | 1.04 | 0.95 | 1.13 | 1.03 | 0.94 | 1.13 |
| Sex, Female vs Male | | |  |  |  | 1.03 | 0.85 | 1.24 | 0.98 | 0.78 | 1.23 | 0.96 | 0.77 | 1.20 | 0.95 | 0.75 | 1.20 | 0.94 | 0.75 | 1.17 |
| Body Mass Index | | |  |  |  | 1.01 | 0.99 | 1.03 | 1.01 | 0.99 | 1.03 | 1.01 | 0.99 | 1.04 | 1.01 | 0.99 | 1.04 | 1.01 | 0.99 | 1.04 |
| Comorbidities, Y vs N | | |  |  |  | 1.13 | 0.93 | 1.37 | 1.18 | 0.99 | 1.41 | 1.15 | 0.92 | 1.42 | 1.10 | 0.87 | 1.40 | 1.10 | 0.90 | 1.35 |
| Smoking status  (vs never smoker) | | Ex-smoker |  |  |  |  |  |  | 1.01 | 0.82 | 1.25 | 1.03 | 0.81 | 1.32 | 0.99 | 0.79 | 1.24 | 1.00 | 0.77 | 1.28 |
|  |  | Smoker |  |  |  |  |  |  | 1.14 | 0.87 | 1.48 | 1.12 | 0.85 | 1.48 | 1.14 | 0.85 | 1.53 | 1.14 | 0.84 | 1.55 |
| Educational level (ref. primary school/no title | | Secondary school |  |  |  |  |  |  | 1.39 | 1.05 | 1.84 | 1.32 | 0.97 | 1.80 | 1.30 | 0.98 | 1.73 | 1.29 | 0.94 | 1.78 |
|  |  | University degree |  |  |  |  |  |  | 1.57 | 1.10 | 2.25 | 1.39 | 0.96 | 1.99 | 1.31 | 0.90 | 1.92 | 1.30 | 0.89 | 1.92 |
| Employment  (ref. Tertiary sector, other) | | Primary / secondary sector |  |  |  |  |  |  | 1.02 | 0.77 | 1.36 | 0.91 | 0.69 | 1.20 | 0.91 | 0.69 | 1.20 | 0.91 | 0.67 | 1.22 |
|  |  | Tertiary sector, housekeeper |  |  |  |  |  |  | 0.75 | 0.49 | 1.14 | 0.66 | 0.43 | 1.01 | 0.60 | 0.36 | 1.01 | 0.60 | 0.35 | 1.03 |
|  |  | Tertiary sector, education |  |  |  |  |  |  | 1.05 | 0.71 | 1.53 | 0.92 | 0.61 | 1.38 | 0.93 | 0.61 | 1.42 | 0.87 | 0.56 | 1.36 |
|  |  | Tertiary sector, healthcare |  |  |  |  |  |  | 2.29 | 1.24 | 4.24 | 1.76 | 0.91 | 3.41 | 1.29 | 0.66 | 2.51 | 1.17 | 0.64 | 2.12 |
|  |  | Tertiary sector, social |  |  |  |  |  |  | 1.35 | 0.74 | 2.47 | 1.48 | 0.82 | 2.67 | 1.26 | 0.71 | 2.24 | 1.25 | 0.68 | 2.30 |
|  |  | Student/unemployed |  |  |  |  |  |  | 1.05 | 0.61 | 1.79 | 0.97 | 0.56 | 1.68 | 0.99 | 0.56 | 1.75 | 0.99 | 0.57 | 1.70 |
|  |  | Retired |  |  |  |  |  |  | 0.63 | 0.45 | 0.88 | 0.55 | 0.38 | 0.78 | 0.52 | 0.36 | 0.76 | 0.50 | 0.34 | 0.73 |
| Crowding index | | |  |  |  |  |  |  | 1.08 | 0.84 | 1.37 | 1.03 | 0.81 | 1.32 | 0.97 | 0.75 | 1.26 | 0.98 | 0.73 | 1.30 |
| Symptoms (Y vs N) | | |  |  |  |  |  |  |  |  |  | 12.64 | 9.72 | 16.44 | 8.63 | 6.42 | 11.61 | 8.26 | 6.04 | 11.31 |
| Contacts with | infected individ. at home, Y vs N | |  |  |  |  |  |  |  |  |  |  |  |  | 7.34 | 3.62 | 14.90 | 7.47 | 3.81 | 14.62 |
|  | infected individ. outside home, Y vs N | |  |  |  |  |  |  |  |  |  |  |  |  | 9.80 | 6.05 | 15.87 | 9.87 | 5.78 | 16.85 |
|  | symptomatic individ. at home, Y vs N | |  |  |  |  |  |  |  |  |  |  |  |  | 1.48 | 0.75 | 2.93 | 1.50 | 0.77 | 2.91 |
|  | sympt. individ. outside home, Y vs N | |  |  |  |  |  |  |  |  |  |  |  |  | 0.94 | 0.47 | 1.86 | 0.94 | 0.48 | 1.86 |
| Vaccination (ref. not vaccinated) | | Ongoing |  |  |  |  |  |  |  |  |  |  |  |  |  |  |  | 1.59 | 1.08 | 2.33 |
|  |  | Completed |  |  |  |  |  |  |  |  |  |  |  |  |  |  |  | 1.50 | 1.04 | 2.18 |

Abbreviations: AIC, Akaike Information Criterion; OR, odds ratio; L95 and U95: lower and upper limits of the 95% confidence interval

**Supplementary Table 6.** Results of mixed-effect logistic regression models adjusted for age, sex, and month, where each displayed variable was included one at a time.

| Variable | OR (95% CI) |
| --- | --- |
| Crowding index | 1.15 (0.92-1.43) |
| Educational qualification (reference Primary school or no title) |  |
| Secondary school | 1.45 (1.10-1.92) |
| University degree | 1.59 (1.13-2.23) |
| Main occupation (reference: Tertiary sector (other)) |  |
| Primary and secondary sector | 1.00 (0.76-1.32) |
| Tertiary sector (housekeeper) | 0.78 (0.49-1.26) |
| Tertiary sector (education) | 1.11 (0.77-1.60) |
| Tertiary sector (healthcare) | 2.47 (1.38-4.41) |
| Tertiary sector (social) | 1.48 (0.84-2.62) |
| Student or unemployed | 1.01 (0.61-1.69) |
| Retired | 0.63 (0.44-0.90) |
| Smoking status (reference: Never smoker) |  |
| Past smoker | 1.04 (0.80-1.34) |
| Current smoker | 1.17 (0.92-1.49) |
| Body Mass Index, *kg/m^2^* | 1.01 (0.99-1.03) |
| Any pathologies (Yes vs No) | 1.14 (0.94-1.38) |
| Symptoms (Yes vs No) | 12.41 (9.42-16.35) |
| Contacts with infected individuals within home (Yes vs No) | 22.71 (13.11-39.12) |
| Contacts with infected individuals outside home (Yes vs No) | 13.47 (9.28-19.55) |
| Contacts with symptomatic individuals within home (Yes vs No) | 10.84 (7.10-16.97) |
| Contacts with symptomatic individuals outside home (Yes vs No) | 8.56 (5.39-13.61) |
| Vaccination status (reference: Not vaccinated) |  |
| Initiated | 2.07 (1.50-2.86) |
| Completed | 1.57 (1.13-2.19) |

*Adjusted for age, sex, and month.

Abbreviations: OR: odds ratio; CI: confidence interval.

**Supplementary Table 7.** Results of the multivariable mixed-effect logistic regression model using m = 3 and m = 50 imputed datasets.

| Variable | m = 3 | m = 50 |
| --- | --- | --- |
|  | OR (95% CI) | OR (95%CI) |
| Age, *per decade* | 1.03 (0.94-1.13) | 1.04 (0.95-1.15) |
| Sex (Female vs Male) | 0.94 (0.75-1.17) | 0.91 (0.71-1.16) |
| Month (reference: September) |  |  |
| October | 0.32 (0.21-0.50) | 0.32 (0.21-0.50) |
| November | 0.65 (0.45-0.92) | 0.64 (0.46-0.91) |
| December | 7.19 (5.35-9.66) | 7.24 (5.33-9.85) |
| January | 0.90 (0.64-1.26) | 0.91 (0.65-1.28) |
| February | 0.97 (0.68-1.38) | 0.98 (0.70-1.38) |
| March | 1.84 (1.36-2.58) | 1.85 (1.31-2.61) |
| April | 1.39 (1.08-1.97) | 1.40 (0.98-2.00) |
| May | 1.53 (1.06-2.20) | 1.53 (1.06-2.21) |
| Crowding index | 0.98 (0.73-1.30) | 0.97(0.74-1.26) |
| Educational qualification (reference Primary school or no title) |  |  |
| Secondary school | 1.29 (0.94-1.78) | 1.31 (0.96-1.78) |
| University degree | 1.30 (0.89-1.92) | 1.29 (0.87-1.93) |
| Main occupation (reference: Tertiary sector (other)) |  |  |
| Primary and secondary sector | 0.91 (0.67-1.22) | 0.91 (0.67-1.23) |
| Tertiary sector (housekeeper) | 0.60 (0.35-1.03) | 0.69 (0.42-1.12) |
| Tertiary sector (education) | 0.87 (0.56-1.36) | 0.92 (0.61-1.39) |
| Tertiary sector (healthcare) | 1.17 (0.64-2.12) | 1.27 (0.72-2.23) |
| Tertiary sector (social) | 1.25 (0.68-2.30) | 1.32 (0.73-2.36) |
| Student or unemployed | 0.99 (0.57-1.70) | 1.04 (0.58-1.86) |
| Retired | 0.50 (0.34-0.73) | 0.46 (0.31-0.68) |
| Smoking status (reference: Never smoker) |  |  |
| Past smoker | 1.00 (0.77-1.28) | 0.97 (0.75-1.26) |
| Current smoker | 1.14 (0.84-1.55) | 1.13 (0.84-1.53) |
| Body Mass Index, *kg/m^2^* | 1.01 (0.99-1.04) | 1.01 (0.99-1.04) |
| Any pathologies (Yes vs No) | 1.10 (0.90-1.35) | 1.12 (0.89-1.39) |
| Symptoms (Yes vs No) | 8.26 (6.04-11.31) | 8.43 (6.30-11.27) |
| Contacts with infected individuals within home (Yes vs No) | 7.47 (3.81-14-62) | 7.31 (3.57-14.98) |
| Contacts with infected individuals outside home (Yes vs No) | 9.87 (5.78-16.85) | 9.92 (5.92-16.61) |
| Contacts with symptomatic individuals within home (Yes vs No) | 1.50 (0.77-2.91) | 1.40 (0.73-2.66) |
| Contacts with symptomatic individuals outside home (Yes vs No) | 0.94 (0.48-1.86) | 0.95 (0.47-1.93) |
| Vaccination status (reference: Not vaccinated) |  |  |
| Initiated | 1.59 (1.08-2.33) | 1.56 (1.04-2.32) |
| Completed | 1.50 (1.04-2.18) | 1.46 (1.02-2.10) |

Abbreviations: OR: odds ratio; CI: confidence interval;

**Supplementary Table 8.** Results of the multivariable mixed-effect logistic regression model using the complete case analysis that included 620 individuals and 3,826 follow-up questionnaires.

| Variable | OR (95% CI) |
| --- | --- |
| Age, *per decade* | 1.06 (0.93-1.21) |
| Sex (Female vs Male) | 0.85 (0.67-1.09) |
| Month (reference: September) |  |
| October | 0.36 (0.22-0.59) |
| November | 0.71 (0.46-1.08) |
| December | 8.27 (5.75-11.90) |
| January | 0.98 (0.68-1.40) |
| February | 1.12 (0.78-1.61) |
| March | 2.30 (1.62-3.29) |
| April | 1.61 (1.05-2.47) |
| May | 1.82 (1.22-2.71) |
| Crowding index | 1.32 (0.97-1.80) |
| Educational qualification (reference Primary school or no title) |  |
| Secondary school | 1.28 (0.93-1.75) |
| University degree | 1.44 (0.89-2.34) |
| Main occupation (reference: Tertiary sector (other)) |  |
| Primary and secondary sector | 1.61 (0.90-2.88) |
| Tertiary sector (housekeeper) | 0.67 (0.36-1.27) |
| Tertiary sector (education) | 1.38 (0.78-2.43) |
| Tertiary sector (healthcare) | 2.22 (1.04-4.74) |
| Tertiary sector (social) | 1.83 (1.05-3.19) |
| Student or unemployed | 2.10 (1.03-4.28) |
| Retired | 1.85 (0.78-4.40) |
| Smoking status (reference: Never smoker) |  |
| Past smoker | 1.15 (0.84-1.57) |
| Current smoker | 1.32 (0.96-1.04) |
| Body Mass Index, *kg/m^2^* | 1.01 (0.98-1.04) |
| Any pathologies (Yes vs No) | 1.12 (0.85-1.48) |
| Symptoms (Yes vs No) | 7.49 (5.58-10.05) |
| Contacts with infected individuals within home (Yes vs No) | 6.81 (3.16-14.67) |
| Contacts with infected individuals outside home (Yes vs No) | 9.37 (4.91-17.90) |
| Contacts with symptomatic individuals within home (Yes vs No) | 1.39 (0.69-2.79) |
| Contacts with symptomatic individuals outside home (Yes vs No) | 0.79 (0.36-1.76) |
| Vaccination status (reference: Not vaccinated) |  |
| Initiated | 1.50 (0.98-2.28) |
| Completed | 1.72 (1.10-2.70) |
| Abbreviations: OR, odds ratio; CI, confidence interval.  Reference categories for categorical variables: see footnote of Supplementary Table 6. | |

**Supplementary Table 9.** Results of multivariable mixed-effect logistic regression model stratified by period.

| Variable | September – December 2020* | January –  May 2021** |
| --- | --- | --- |
|  | OR (95% CI) | OR (95%CI) |
| Age, *per decade* | 1.10 (0.98-1.23) | 0.97 (0.86-1.11) |
| Sex (Female vs Male) | 0.98 (0.72-1.35) | 0.90 (0.63-1.29) |
| Crowding index | 0.91 (0.65-1.28) | 0.98 (0.70-1.37) |
| Educational level (reference: Primary school or no title) |  |  |
| Secondary school | 1.14 (0.76-1.70) | 1.40 (0.95-2.07) |
| University degree | 1.16 (0.69-1.93) | 1.44 (0.89-2.35) |
| Main occupation (reference: Tertiary sector (other)) |  |  |
| Primary and secondary sector | 0.94 (0.63-1.42) | 0.94 (0.64-1.38) |
| Tertiary sector – housekeeping | 0.52 (0.26-1.04) | 0.75 (0.41-1.38) |
| Tertiary sector – education | 1.13 (0.65-1.96) | 0.72 (0.41-1.38) |
| Tertiary sector – healthcare | 1.15 (0.53-2.48) | 1.04 (0.44-2.49) |
| Tertiary sector – social | 1.48 (0.65-3.36) | 1.13 (0.56-2.30) |
| Student or unemployed | 1.27 (0.54-3.01) | 0.87 (0.39-1.92) |
| Retiree | 0.46 (0.28-0.75) | 0.51 (0.30-0.87) |
| Smoking status (reference: Never smoker) |  |  |
| Past smoker | 1.18 (0.83-1.67) | 0.86 (0.60-1.22) |
| Current Smoker | 1.17 (0.82-1.68) | 1.11 (0.76-1.63) |
| Body Mass Index, *kg/m^2^* | 1.03 (1.00-1.06) | 1.00 (0.97-1.04) |
| Any pathologies (Yes vs No) | 1.16 (0.90-1.35) | 1.07 (0.81-1.41) |
| Any reported symptoms (Yes vs No) | 6.91 (4.37-10.94) | 10.42 (7.11-15.26) |
| Contacts with infected individuals within home (Yes vs No) | 9.98 (3.81-26.13) | 4.30 (1.69-10.94) |
| Contacts with infected individuals outside home (Yes vs No) | 7.45 (3.87-14.35) | 14.65 (6.71-32.00) |
| Contacts with symptomatic individuals within home (Yes vs No) | 0.80 (0.31-2.07) | 3.98 (1.37-11.57) |
| Contacts with symptomatic individuals outside home (Yes vs No) | 1.30 (0.59-2.86) | 0.65 (0.22-1.94) |
| Vaccination status (reference.: Not vaccinated) |  |  |
| Incomplete | NA | 1.81 (1.25-2.62) |
| Complete | NA | 1.62 (1.08-2.45) |
| Abbreviations: OR: odds ratio; CI: confidence interval; NA: not applicable.  *Adjusted for month: the ORs for October, November and December using September as the reference month were of 0.34 (95%CI: 0.21-0.54), 0.70 (95%CI: 0.47-1.03), and 6.93 (95%CI: 4.92-9.76), respectively.  ** Adjusted for month: the ORs of February, March, April, and May 2021, using January as the reference month were of 1.04 (95%CI: 0.75-1.41), 2.15 (95%CI: 1.59-2.92), 1.57 (95%CI: 1.14-2.16), and 1.72 (95%CI: 1.23-2.42), respectively. | | |

**Supplementary Figure 1.** Relative frequencies of missing values for the predictors included in the analyses.

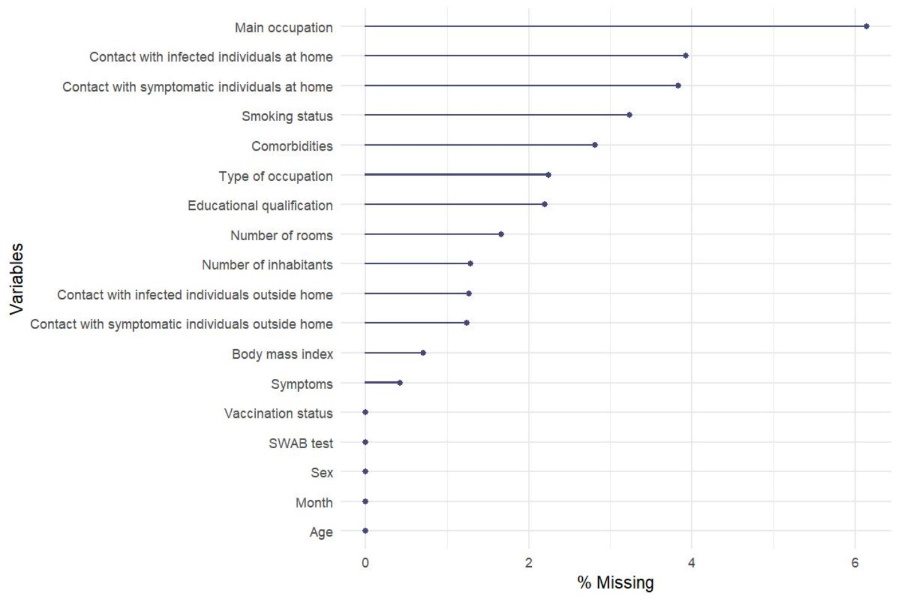

**Supplementary Figure 2.** Missing data pattern. The lower-left panel shows the absolute count of missing values per variable. The lower-right panel shows the most frequent combinations of missing values among variables. The upper-right panel displays the absolute frequencies of missing values corresponding to each combination.

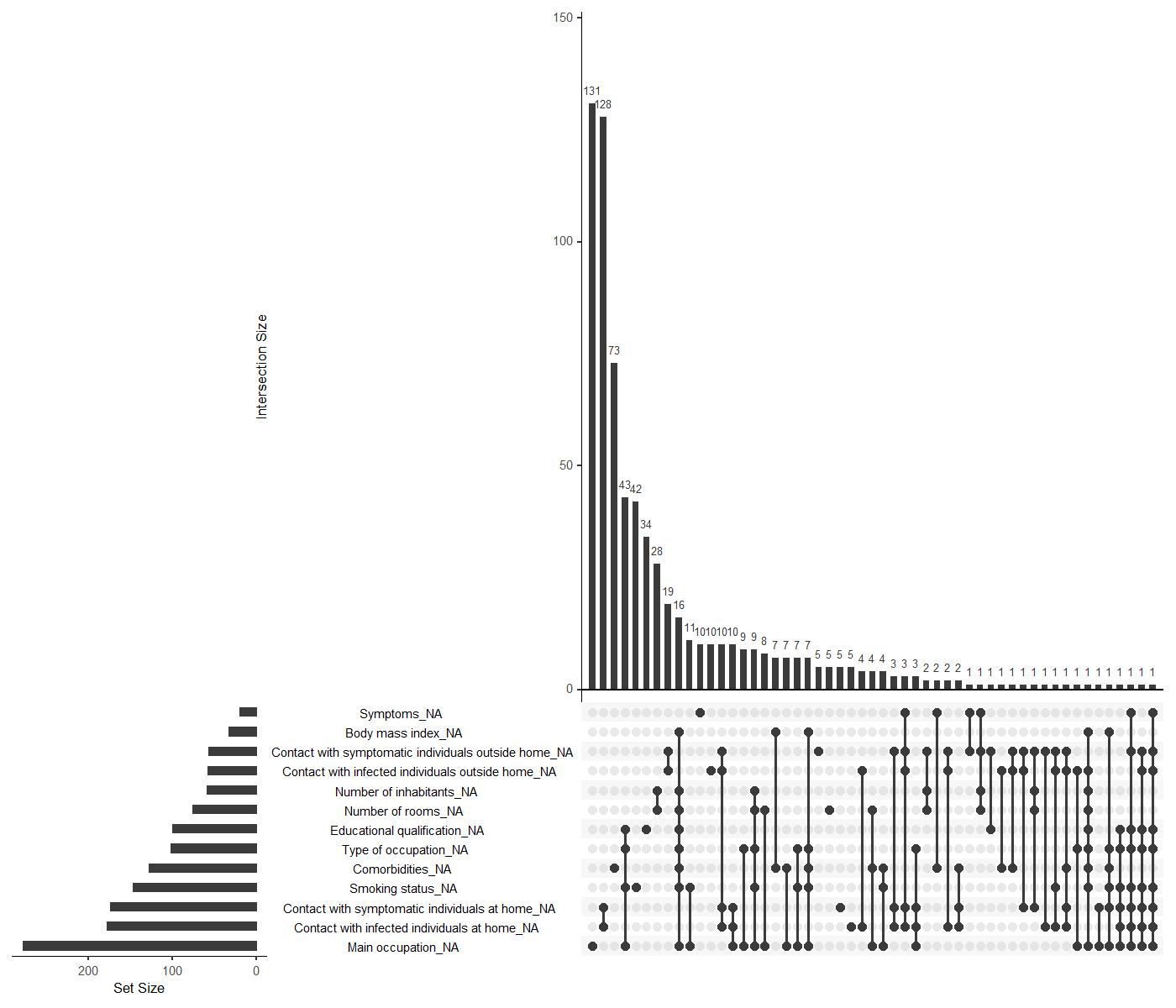

**Supplementary Figure 3.** Convergence analysis of the MICE procedure for the variable ‘Main Occupation’. **Panel A**: trace plots using 3 imputed datasets (50 iterations). The top plot indicates the mean of the imputed (not observed) values of Main Occupation across sequential iterations for every imputed data (each line represents one of the three imputed datasets). The bottom plot represents the corresponding standard deviation of the imputed (not observed) values of Main Occupation. **Panel B**: trace plots using 50 imputed datasets (50 iterations).

**A**

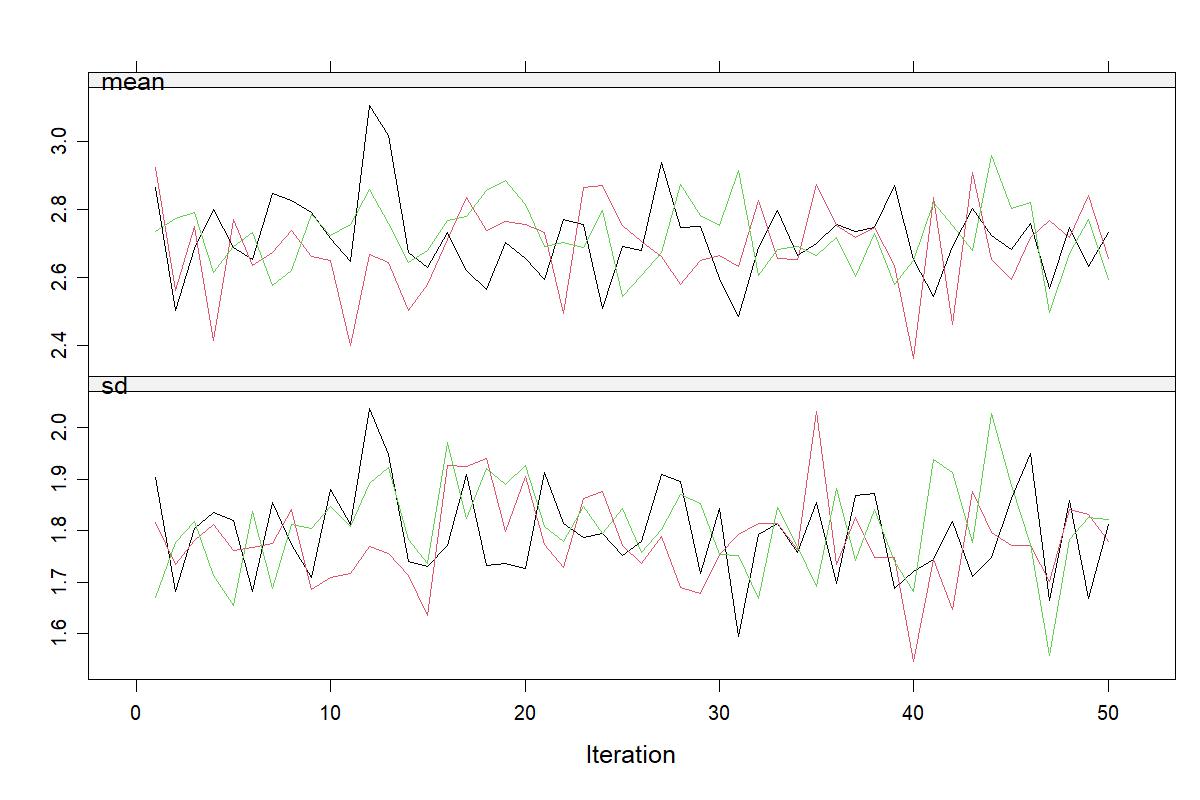

**B**

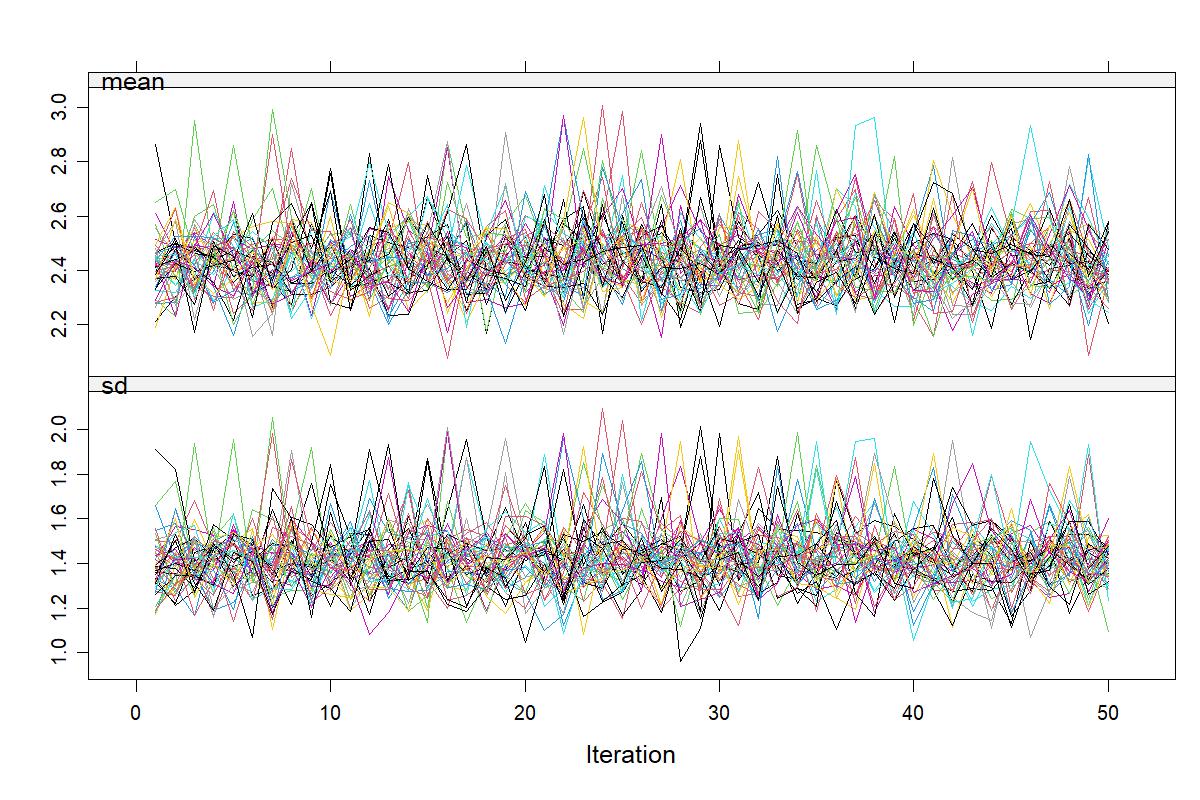
